# Data Resource Profile: thousands of circulating RNA profiles of pre-clinical samples from the Janus Serum Bank Cohort

**DOI:** 10.1101/2021.01.22.20243154

**Authors:** Hilde Langseth, Sinan Ugur Umu, Cecilie Bucher-Johannessen, Ronnie Babigumira, Magnus Leithaug, Marianne Lauritzen, Paolo Vineis, Giske Ursin, Robert Lyle, Trine B Rounge

## Abstract

There is justified optimism regarding the use of miRNAs as early detection biomarkers of cancer. They are well characterized and are involved in all the hallmarks of cancer. Less is known about the role of most other non-coding RNA (ncRNAs) classes in normal physiology and tumorigenesis. The JanusRNA dataset consist of circulating RNA profiles of pre-clinical samples from 1631 cancer patients and 673 cancer-free controls. We studied eight cancer types including cancer of the: lung, colon, rectum, prostate, breast, testis, ovaries and gallbladder. JanusRNA has its origin from the large population-based Janus Serum Bank Cohort which consists of 318 628 Norwegians. The dataset combines information from the complete nationwide cancer registry, RNA sequencing profiles from 1631 cancer patients and 673 cancer-free controls, as well as data on lifestyle, anthropometry and biochemical measurements from national health surveys. The Janus Serum Bank is specifically suited for studies of early detection and risk biomarkers of cancer, since samples are collected nationwide over a large time span, pre-clinically and cancer occurs at different points in time after blood draw. We used a nested case-control design, selecting both cases and controls among the Janus cohort members. We restricted our selection to cases with at least one sample collected within 10 years prior to cancer diagnosis. We selected 673 cancer-free Janus participants for comparison of RNA levels with the cancer cases. The controls were frequency matched to the case group on sex, age at blood donation and date of blood donation. The JanusRNA dataset has been used to investigate the natural variation of circulating RNAs in cancer-free individuals. This data resource was also used in a study of variation in RNA expression associated with common traits like age, sex, smoking, BMI and physical activity in cancer-free individuals. RNA dynamics in lung and testicular carcinogenesis throughout a 10-year follow-up has also been studied.

## Data resource basics

There is justified optimism regarding the use of miRNAs as early detection biomarkers of cancer. They are well characterized and are involved in all the hallmarks of cancer (1). Less is known about the role of most other non-coding RNA (ncRNAs) classes in normal physiology and tumorigenesis. Several classes of ncRNA act as regulators of key cellular processes, many of which are associated with cancer (2). Circulating ncRNAs may improve cancer management in the future as minimally invasive biomarkers. Large prospective cohorts with harmonized genetic and phenotypic data are required to realize the potential of ncRNAs as early cancer biomarkers, but such datasets are rare (3).

In this resource profile, we present the circulating RNA dataset available within the large prospective Janus Serum Bank Cohort in Norway (Janus RNA). The resource was established to facilitate research on RNA dynamics in samples collected up to 10 years prior to cancer diagnosis, compared to cancer-free controls, and with the long-term objective of identifying early detection biomarkers of cancer. The data resource will be instrumental for a wide range of national and international cancer research projects in the future.

We studied eight cancer types. Five of these cancer types are of major public health concern in many countries: lung cancer (LC), colon cancer (CC), rectum cancer (REC), prostate (PC) and breast cancer (BC). In all of them early detection screening biomarkers could have big public health implications. We included testicular germ cell tumors (TGCT), the most common malignancy in young males in most Western countries. Further, we expanded the dataset with cancer of the ovaries (OC) and gallbladder (GBC) which are diseases with poor prognosis since they are often diagnosed at advanced stages.

## Data collected

JanusRNA has its origin in the large population-based Janus Serum Bank Cohort which consists of 318 628 Norwegians. The serum bank is administered by the Cancer Registry of Norway (CRN) (4). The dataset combines information from the complete nationwide cancer registry (5), RNA sequencing profiles from 1631 cancer patients and 673 cancer-free controls, as well as data on lifestyle, anthropometry and biochemical measurements from national health surveys (6) (Figure 1). The Janus Serum Bank is specifically suited for studies of early detection and risk biomarkers, since samples were collected nationwide over a large time span, pre-clinically. Cancer has occurred at different points in time after the blood draw.

**Figure 1.**
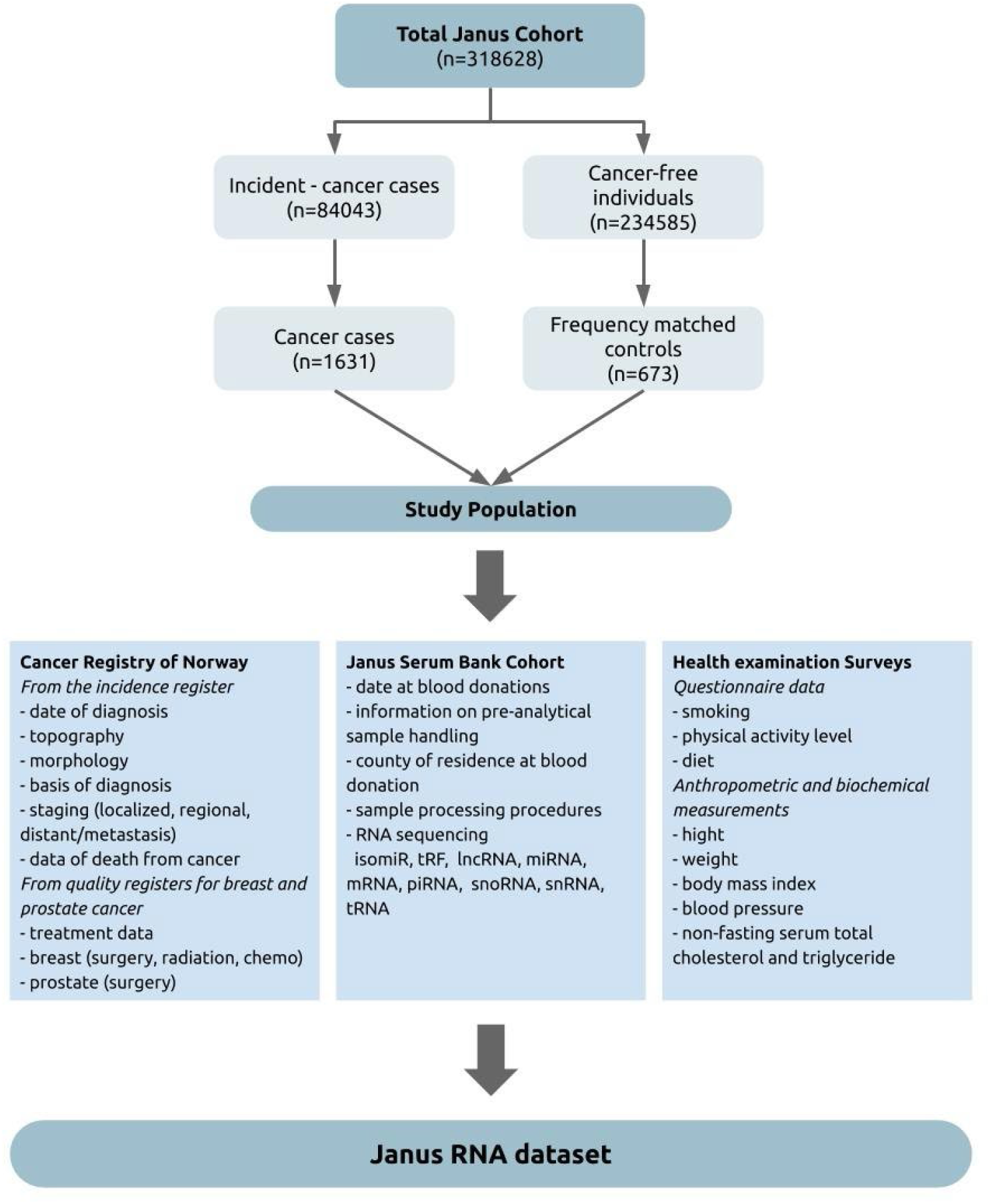
Study population and data sources.

## Study design and Sampling strategy

We used a nested case-control design, selecting both cases and controls among the Janus cohort members. This design offers logistic efficiency, is typically used for molecular epidemiological studies within prospective cohorts, and is suited for studying biomarkers that can be influenced by analytical batch, long-term storage and freeze-thaw cycles (7).

### Case selection

The cases were identified by linking the Janus Cohort to the CRN using the individual’s Norwegian national identity number. We included cohort participants with the selected cancer sites as their primary cancer diagnosis. No prior cancer diagnosis (except non-melanoma skin cancer) were allowed, and at least 500 µl of serum had to be available. We restricted our selection to cases with at least one sample collected within 10 years prior to cancer diagnosis. The dynamics in RNA levels prior to a cancer diagnosis is sparsely described, however studies in Janus showed that most changes in RNA expression occurred close to a lung cancer diagnosis (8), and depend on staging (9). Based on our knowledge from using pre-diagnostic samples we considered 10 years prior to diagnosis as a reasonable timeframe for the study objectives. The average age of recruitment to the Janus cohort is 41 years. Because most cancers occur at older ages, most of the eligible cancer cases have a long lag-time between sampling and diagnosis. Therefore, we selected all available cases in the time window up to five years prior to diagnosis and a selection of eligible cases in the 5-10 years prior to diagnosis. Distribution of case samples in the different pre-diagnostic time-windows are shown in Figure 2.

**Figure 2.**
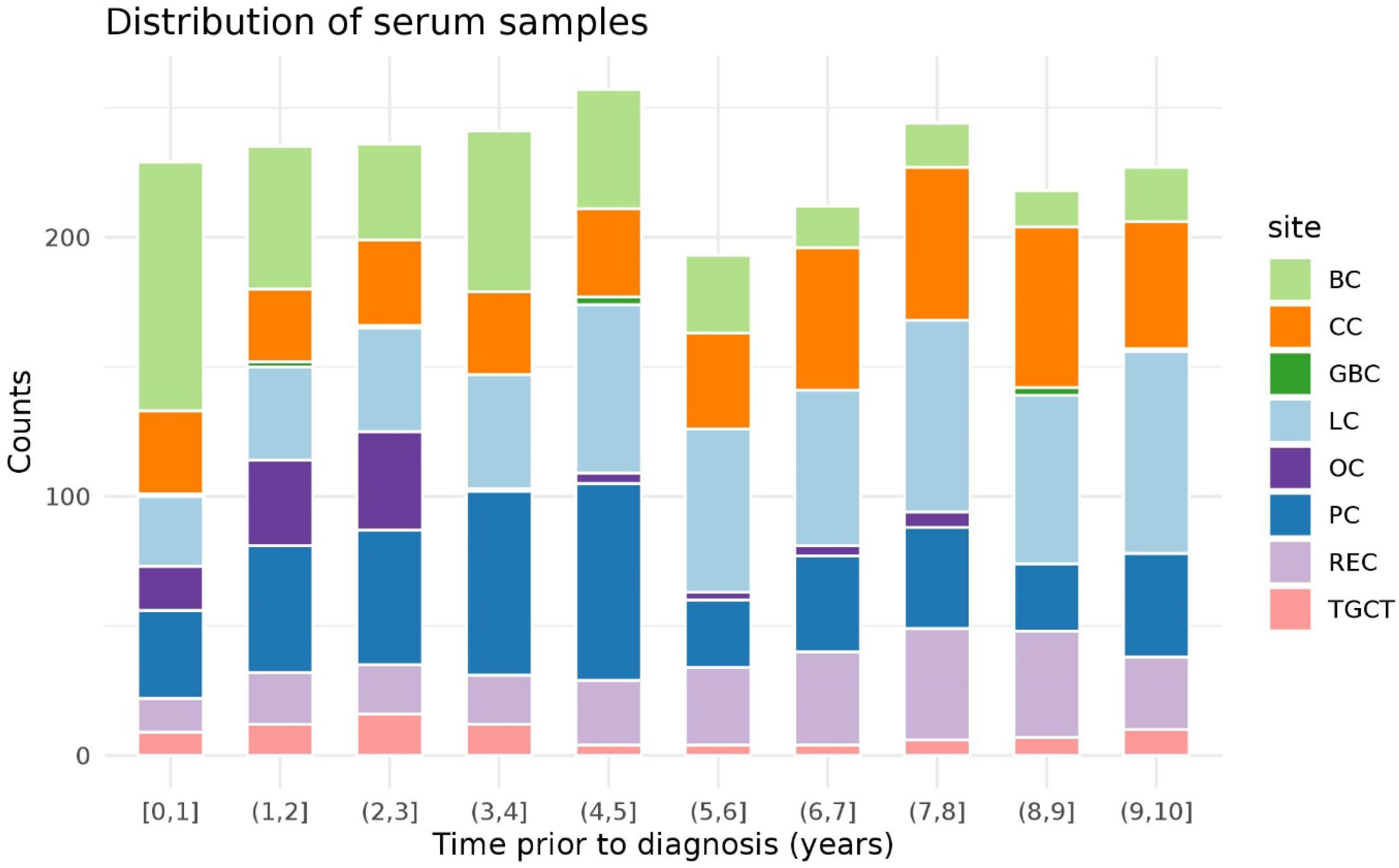
Sample distribution by pre-diagnostic collection timepoints. One year time window from 0 to 10 years prior to diagnosis on the x-axis and number of samples by cancer type on the y-axis.

### Control selection

Controls were selected according to a modified version of the incidence density sampling method for nested case-control studies (10). The cases were set up in strata based on age at blood sampling, gender, time period and county of residence at sampling. The date of diagnosis for the latest case in the case strata, was set as limit for all controls in the strata. The controls had to be alive and free from cancer at this date to be included in the control-pool. In addition, they had to be free from cancer up to ten year after blood collection. Due to the many unknown factors that affect the RNA levels prior to a cancer diagnosis, we considered it appropriate to set these additional criteria for the controls to give the best basis of comparisons between cases and controls. We then randomly selected frequency matched controls such that the case/control ratio was the same within each stratum (11).

We profiled circulating RNA by sequencing 2 997 serum samples from 1 631 cancer cases. This includes cancer of the lung (n=404), colon (n=308), rectum (n=182), breast (n=206), prostate (n=332), testis (n=84), gallbladder (n=27) and ovaries (n=88). Gender, age at blood donation and age at diagnosis by cancer type is shown in Table 1. There are 199 cases with a second cancer diagnosis and 22 cases with a third cancer diagnosis. Furthermore, 107 of the controls developed a cancer diagnosis 10 years or more after the blood donation (not shown in table). Multiple samples (up to 7 time points) are available for a subset of the cancer patients, allowing investigation of temporal variations in samples from the same individual (Table 1). RNA profiles were also produced from 673 cancer-free control samples collected at a single time point. A pooled positive control sample, consisting of serum from several individuals, was profiled 28 times for quality and reproducibility assessment. In addition, for each batch of 96 samples, we included two negative control samples, one negative extraction control (NEC) and one water control from the library preparations.

**Table 1.** Basic characteristics of the study subjects and samples included in the JanusRNA dataset, by gender.

| Cancer site ICD 10 | Men (n=1379) |  |  |  |  |  |  |  | Women (n=925) |  |  |  |  |  |  |  |
| --- | --- | --- | --- | --- | --- | --- | --- | --- | --- | --- | --- | --- | --- | --- | --- | --- |
|  | Lung<br>(C33-34) | Colon<br>(C18) | Rectum<br>(C19-20) | Gall<br>bladder<br>(C23) | Prostate<br>(C61) | Testis<br>(C62) | Contr<br>- | TOT | Lung<br>(C33-34) | Colon<br>(C18) | Rectum<br>(C19-20) | Gall<br>bladder<br>(C23) | Breast<br>(C50) | Ovary<br>(C56) | Contr<br>- | TOT |
| No of individuals | 274 | 162 | 118 | 7 | 332 | 84 | 402 | 1379 | 130 | 146 | 64 | 20 | 206 | 88 | 271 | 925 |
| Age at recruitment | 54 (8.61) | 53.1 (9.5) | 52.9 (8.19) | 44.1 (3.28) | 59.9 (8.67) | 34.8 (6.46) | 50.7 (12) | - | 53.4 (8.5) | 53.2 (9.92) | 51.2 (8.53) | 47.3 (8.87) | 45.5 (6.97) | 46.7 (8.57) | 48.5 (10.2) | - |
| Age at diagnosis, mean (SD) | 60.2 (8.58) | 59 (9.19) | 59.3 (8.09) | 53.4 (3.15) | 64.5 (7.99) | 38.6 (5.07) | 74 (9.12) | - | 59.3 (8.13) | 59.2 (9.64) | 57.7 (8.62) | 56.4 (10) | 49.3 (7.68) | 48.8 (8.60) | 70.8 (8.29) | - |
| Total # of samples | 382 | 231 | 183 | 7 | 450 | 84 | 405 | 1742 | 177 | 193 | 93 | 20 | 395 | 106 | 271 | 1255 |
| No with 1 sample | 270 | 162 | 118 | 7 | 331 | 84 | 402 | 1374 | 130 | 144 | 64 | 20 | 206 | 88 | 271 | 923 |
| No with 2 samples | 77 | 46 | 41 | 0 | 68 | 0 | 3 | 235 | 40 | 42 | 24 | 0 | 132 | 14 | 0 | 252 |
| No with $\leq 3$ samples | 35 | 23 | 24 | 0 | 51 | 0 | 0 | 133 | 7 | 7 | 5 | 0 | 57 | 4 | 0 | 80 |

## Data set production

JanusRNA contains harmonized data from three sources: clinical cancer records from CRN, lifestyle information from nationwide health surveys, and RNA profiles and sampling information from Janus (Figure 1).

### Cancer Registry data

Since1952, CRN has systematically collected mandatory notifications on cancer occurrence for the Norwegian population. The registration is considered to be close to complete from 1953, with 98.8% completeness for the registration period 2001– 2005 (5). Information from clinical notifications, pathology reports and death certificates are the main sources that enables the CRN to code and store data on cancer patients in Norway. Information from the Norwegian Patient Registry (NPR) is an important additional source for identifying cancer cases. Clinical registries, also administered by CRN provide detailed information about diagnostic procedures, pathology-examinations, treatment and follow-up from cases that were diagnosed after 2004 (12).

### Health survey data on lifestyle, anthropometry and biochemical measurements

Survey data were collected at the time of blood donation from most study participants (Table 2). The quality, completeness and standardization of these data has been described elsewhere (6). In brief, participants in the health surveys completed a baseline questionnaire about smoking habits and physical activity, and anthropometry and blood pressure were measured. Biochemical parameters such as cholesterol, triglycerides and glucose were also measured. As shown in (Table 2), the proportion of current smokers ranges from 23% in rectal cancer to 70% in lung cancer in males, and from 27% in breast cancer to 70% in lung cancer in females. The percentage of obese individuals (BMI ≥ 30kg/m^2^) ranges from 2% in testicular cancer cases to 14% in male colon cancer cases, in females BMI ranges from 8% in breast cancer cases to 20% in gallbladder cancer cases. Sedentary physical activity levels (inactive + low) varied from 43% in male gall bladder cases to 69% in male lung cancer cases. In women, 60% of the breast cancer cases and 87% of the ovarian cancer cases reported sedentary physical activity habits.

**Table 2.** Selected health survey data for the JanusRNA participants

| Cancer site ICD 10 | Men (n=1376) |  |  |  |  |  |  | Women (n=934) |  |  |  |  |  |  |
| --- | --- | --- | --- | --- | --- | --- | --- | --- | --- | --- | --- | --- | --- | --- |
|  | Lung<br>(C33-34) | Colon<br>(C18) | Rectum<br>(C19-20) | Gall<br>bladder<br>(C23) | Prostate<br>(C61) | Testis<br>(C62) | Contr<br>- | Lung<br>(C33-34) | Colon<br>(C18) | Rectum<br>(C19-20) | Gall<br>bladder<br>(C23) | Breast<br>(C50) | Ovary<br>(C56) | Contr<br>- |
| **Smoking n (%)** | 239 (87) | 135 (83) | 88 (75) | 7 (100) | 265 (80) | 63 (75) | 287 (72) | 114 (88) | 131 (90) | 57 (89) | 20 (100) | 141 (68) | 88 (94) | 183 (68) |
| Current | 191 (70) | 45 (28) | 27 (23) | 3 (43) | 91 (27) | 25 (29) | 105 (26) | 91 (70) | 53 (36) | 20 (31) | 6 (30) | 55 (27) | 33 (37) | 67 (25) |
| Former | 46 (16) | 61 (37) | 44 (38) | 3 (43) | 118 (36) | 19 (23) | 108 (27) | 20 (16) | 33 (23) | 6 (9) | 4 (20) | 24 (12) | 15 (17) | 39 (14) |
| Never | 2 (1) | 29 (18) | 17 (14) | 1 (14) | 56 (17) | 19 (23) | 77 (19) | 3 (2) | 45 (31) | 31 (49) | 10 (50) | 62 (30) | 35 (40) | 77 (29) |
| Missing | 35 (13) | 27 (17) | 30 (25) | 0 (0) | 67 (20) | 21 (25) | 112 (28) | 16 (12) | 15 (10) | 7 (11) | 0 (0) | 65 (31) | 5 (6) | 88 (32) |
| **Body mass index (mean and SD)** | 25.1 (3.59) | 26.4 (3.63) | 26.3 (3.52) | 24.6 (1.63) | 26.3 (3.55) | 25.2 (3.52) | 25.1 (2.9) | 25.2 (4.96) | 25.9 (4.57) | 25.8 (4.51) | 26.8 (5.47) | 24.7 (4.34) | 24.4 (3.75) | 24.9 (3.75) |
| **Body mass index n (%)** | 239 (87) | 135 (83) | 88 (75) | 7 (100) | 265 (80) | 62 (74) | 72 | 114 (88) | 130 (89) | 57 (89) | 20 (100) | 142 (69) | 84 (95) | 183 (68) |
| Underweight | 6 (2) | 1 (1) | 1 (1) | 0 (0) | 3 (1) | 0 (0) | 1 (0.2) | 3 (2) | 1 (1) | 3 (5) | 0 (0) | 0 (0) | 3 (3) | 4 (1) |
| Normal | 113 (41) | 51 (31) | 35 (30) | 4 (57) | 99 (30) | 29 (35) | 135 (33) | 62 (48) | 66 (45) | 27 (42) | 11 (55) | 93 (45) | 48 (55) | 86 (32) |
| Overweight | 102 (37) | 61 (38) | 37 (31) | 3 (43) | 124 (37) | 31 (37) | 132 (33) | 29 (22) | 37 (25) | 20 (31) | 5 (25) | 34 (16) | 25 (28) | 68 (25) |
| Obese | 18 (7) | 22 (14) | 15 (13) | 0 (0) | 39 (12) | 2 (2) | 23 (6) | 20 (16) | 26 (18) | 7 (11) | 4 (20) | 15 (8) | 8 (9) | 26 (10) |
| Missing | 35 (13) | 27 (17) | 30 (25) | 0 (0) | 67 (20) | 22 (26) | 111 (28) | 16 (12) | 16 (11) | 7 (11) | 0 (0) | 64 (31) | 4 (5) | 87 (32) |
| **Physical activity n (%)** | 239 (87) | 133 (82) | 88 (75) | 7 (100) | 257 (77) | 63 (75) | 286 (71) | 114 (88) | 130 (89) | 57 (89) | 19 (95) | 141 (68) | 84 (95) | 181 (67) |
| inactive | 66 (24) | 22 (14) | 24 (20) | 2 (29) | 38 (11) | 16 (19) | 41 (10) | 26 (20) | 25 (17) | 11 (17) | 6 (30) | 28 (13) | 20 (22) | 27 (10) |
| low | 122 (45) | 81 (50) | 47 (40) | 1 (14) | 168 (51) | 32 (38) | 176 (44) | 80 (62) | 94 (64) | 40 (63) | 11 (55) | 97 (47) | 57 (65) | 138 (51) |
| medium | 49 (17) | 28 (17) | 13 (11) | 4 (57) | 51 (15) | 14 (17) | 67 (17) | 8 (6) | 10 (7) | 6 (9) | 2 (10) | 16 (8) | 6 (7) | 14 (5) |
| high | 2 (1) | 2 (1) | 4 (4) | 0 (0) | 0 (0) | 1 (1) | 2 (0.5) | 0 (0) | 1 (1) | 0 (0) | 0 (0) | 0 (0) | 1 (1) | 2 (1) |
| Missing | 35 (13) | 29 (18) | 30 (25) | 0 (0) | 75 (23) | 21 (25) | 116 (29) | 16 (12) | 16 (11) | 7 (11) | 1 (5) | 65 (32) | 4 (5) | 90 (33) |
| **Total cholesterol, mean (SD)** | 6.27 (1.26) | 6.32 (1.05) | 6.47 (1.26) | 5.69 (1.29) | 6.13 (1.17) | 5.71 (1.10) | 6.24 (1.14) | 6.65 (1.36) | 6.62 (1.27) | 6.85 (1.37) | 6.04 (0.82) | 5.86 (1.15) | 6.27 (1.56) | 6.27 (1.33) |
| **Triglycerides, mean (SD)** | 2.14 (1.99) | 2.21 (1.31) | 2.35 (1.86) | 2.43 (1.29) | 2.06 (1.26) | 1.75 (0.98) | 2.15 (1.26) | 1.73 (0.89) | 1.71 (1.43) | 1.91 (1.14) | 1.54 (0.80) | 1.33 (0.77) | 1.40 (0.73) | 1.47 (0.73) |
| **Glucose, mean (SD)** | 5.97 (1.42) | 5.67 (1.06) | 5.82 (1.18) | 6.16 (0.59) | 5.72 (1.11) | 5.27 (0.81) | 5.88 (1.71) | 6.07 (1.82) | 5.78 (1.3) | 5.75 (0.91) | 5.64 (0.60) | 5.81 (0.91) | 5.51 (0.69) | 5.75 (1.09) |

### Sequencing data production

We have developed a platform for small RNA sequencing, tailored to low RNA yield samples such as archived serum samples. This platform enables expression profiling of RNAs from 17-47 nucleotides, annotation of 9 RNA classes and sequence isoform identification. The quality and quantity of RNA was sufficient for analyses, independent of storage time and sample pre-processing (13). Sequencing data production entails three steps:

#### Step 1 – RNA isolation

RNA was extracted from 2 × 200 µl serum using the miRNeasy Serum/Plasma kit (Cat. no 1071073, Qiagen) on a QIAcube (Qiagen). Internal spike-in control *C. elegans* miR-39 was added to each sample. Glycogen (Cat. no AM9510, Invitrogen) was used as carrier during the RNA extraction step. The eluate was concentrated using Ampure beads XP (Agencourt). Cases and controls were blinded.

#### Step 2 - Library preparation

Small RNAseq libraries were created using NEBNext® Small RNA Library Prep Set for Illumina (Cat. No E7300, New England Biolabs Inc.) with a cut size on the Pippin Prep (Cat. No CSD3010, Sage Science) of 17-47 nucleotides.

#### Step 3 - RNA sequencing

12 samples were sequenced per lane on an Illumina HiSeq 2500 platform to an average depth of 18 million reads per sample. A detailed description of the RNA profiles is available (14).

### Bioinformatics pipeline

A custom RNA analysis pipeline for processing raw RNA sequences to count data and expression profiles has been established (Figure 3). It includes adaptor and low-quality data filtering, mapping, and counting of RNA annotations. The RNAseq reads were initially trimmed for adapters using AdapterRemoval (v2.1.7) (15). We then mapped the collapsed reads (generated by FASTX v0.14) to the human genome (hg38) using Bowtie2 (10 alignments per read were allowed). All multi-mapped reads with equivalent mapping score were counted. We compiled a comprehensive annotation set from miRBase (v21) (16) for miRNAs, pirBAse (v1.0) for piRNAs63, GENCODE (v26) (17) for other RNAs and tRNAs. We used SeqBuster (v3.1) (18) to profile isomiR and miRNA profiles. To count the mapped reads, HTSeq (v0.7.2) (19) was used. Candidate tRNA fragments (tRFs) were selected from reads mapped to tRNA annotations. We later updated our workflow and tRFs were profiled using MINTmap tool (https://www.nature.com/articles/srep41184). When reporting the identified RNAs, we excluded RNAs with fewer than 10 reads in more than 20% of the samples (20).

**Figure 3.**
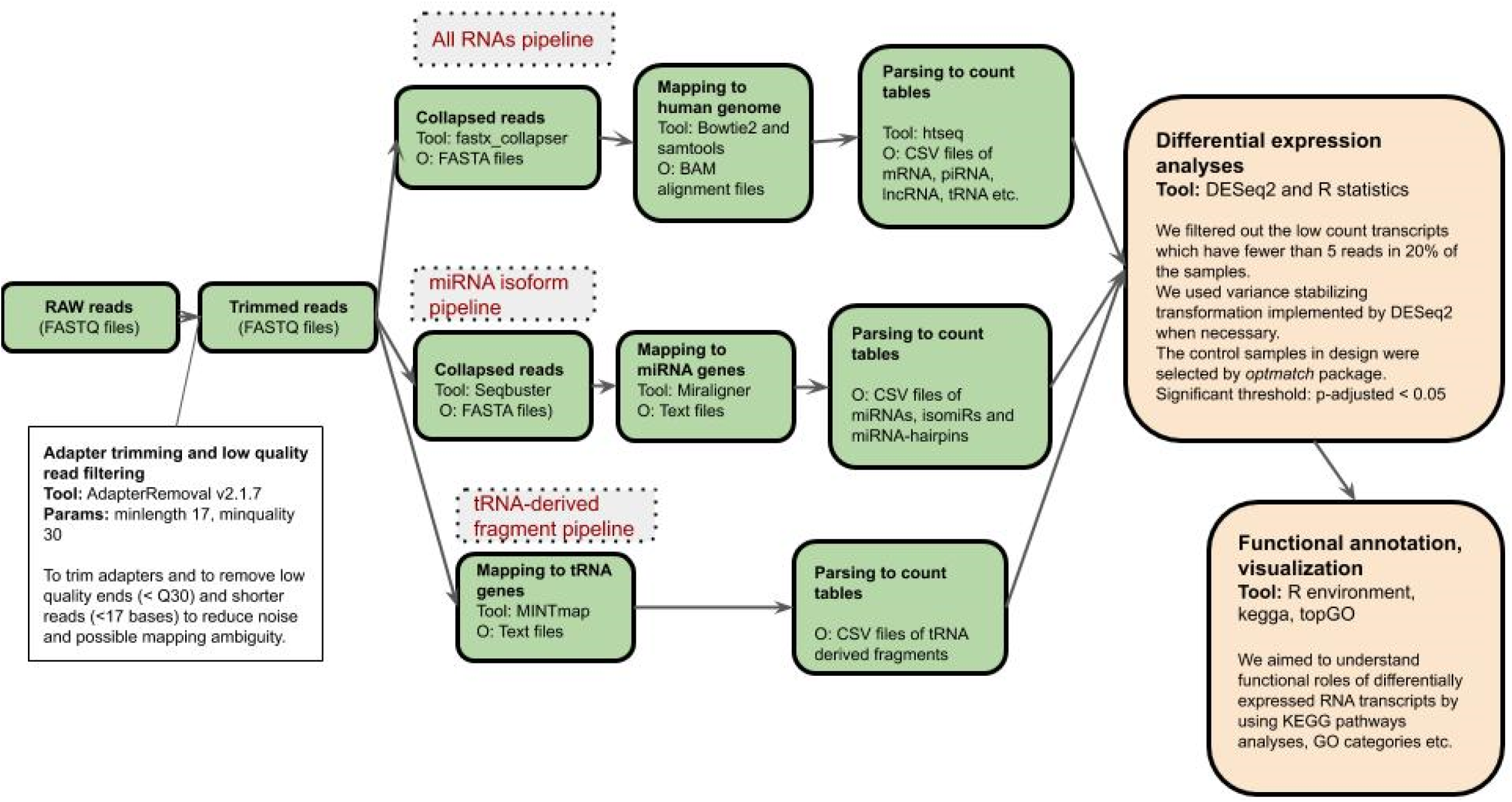
Illustration of the bioinformatic pipeline for all RNA classes identified in JanusRNA, including adaptor and low-quality data clean-up, mapping to the genome, and counting all RNAs between 17 and 47 nucleotides in length.

The average sequencing depth and average number of RNAs by cancer type is given in Figure 4A and Figure 4B. The most abundant RNA type across all cancer types and controls is mRNA followed by isomiRs. The heat-map in figure 4 B shows the average number of identified RNAs for the different cancer types. The RNAseq read counts in millions for all RNA classes combined, ranged from 13.5 in ovarian cancer patients to 20.7 in female gallbladder patients, and standard deviations ranged between 2.9 and 6.2 (Supplementary Table 1).

**Figure 4.**
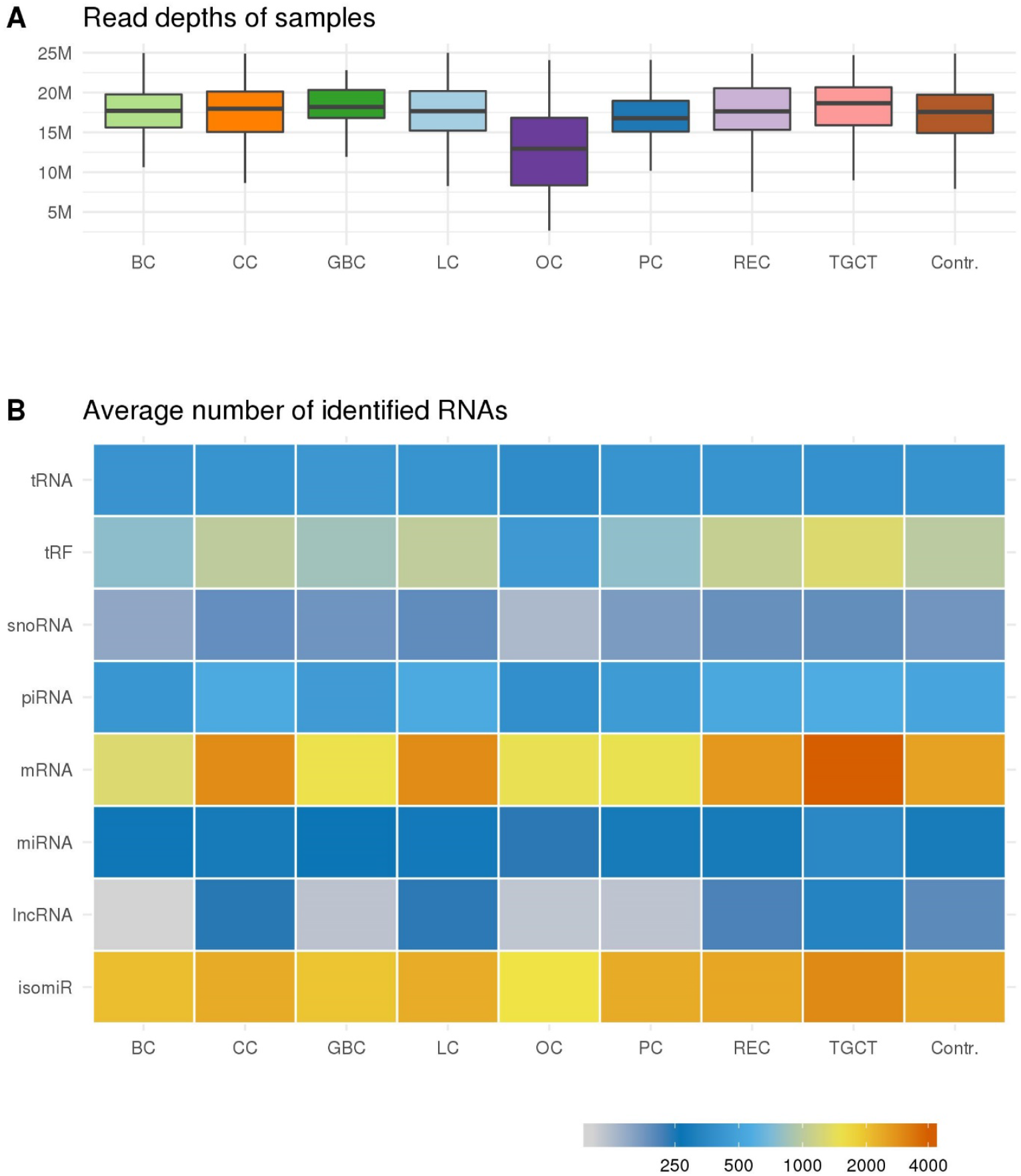
A) The average sequencing depth range from 13.5 mill reads in the pre-diagnostic samples from ovarian cancer patients (OC) to 21 mill reads in gall bladder cancer patients (GBC). B) The heat-map shows the average number of identified RNAs by cancer type. The most highly expressed RNAs are indicated by red color and the lowest expressed by gray color.

## Ethical approval

The JanusRNA study was approved by the regional committee for medical and health research ethics, Oslo, Norway (2016/1290) and (2013/1821), and is based on a broad consent from participants in the Janus cohort. The JanusRNA database contains pseudonymized data.

## Data resource use

The JanusRNA dataset has been used to investigate the natural variation of circulating RNAs in healthy controls (14). The core serum RNA repertoire in the cancer-free control group includes 258 micro RNAs (miRNA), 441 piwi-interacting RNAs (piRNA), 411 transfer RNAs (tRNA), 24 small nucleolar RNAs (snoRNA), 125 small nuclear RNAs (snRNA) and 123 miscellaneous RNAs (misc-RNA). We investigated biological and technical variation in expression, and the results suggest that many RNA molecules identified in serum shows signs of inter-individual variation (14). The same dataset was used to investigate the association between circulating RNAs and common traits such as age, sex, smoking, BMI and physical activity. The study showed that common traits influence circulating RNA expression, in particular age and sex, and concluded that these traits should be treated as potential confounders for RNA analyses (20). RNA dynamics in lung and testicular carcinogenesis throughout a 10-year follow-up has been studied (9, 21). Ongoing studies include pre-diagnostic RNA dynamics in cancer of the lung, colorectal, prostate, breast, gallbladder, testis, ovary and a pan-cancer profile (https://www.kreftregisteret.no/en/Research/Projects/Small-non-coding-RNA-as-early-detection-cancer-biomarkers/).

## Strengths and weaknesses

A strength of JanusRNA is the large number of population-based samples sequenced from eight different cancer types combined with high-quality cancer registry data and access to information on life-style factors at baseline such as BMI, smoking and physical activity, that may influence circulating RNA expression levels significantly. The selection of cases and controls is based on established procedures for linking biobank and registry data (22). We have created a robust and unique dataset to investigate RNA dynamics and biomarker potential across cancer types. It is also a strength that we have sequenced samples from a large cancer-free control group enabling us to study the variation in a healthy population. The control group gives the opportunity to explore disease versus trait-specific patterns, which is important for early detection biomarkers discovery. The control group profiles can be re-used to contrast compatible RNA profiles from a range of clinical studies. Another strength of the JanusRNA dataset is the pre-diagnostic sample collection, which provides the possibility of studying RNA levels prior to diagnosis and compare this with the healthy control group. We also use well-established sequencing platforms, biocomputational capacity and the RNA yield of our biobanked material as documented (13, 14, 20, 23). The sequencing read-depth is high (on average 18 million reads per sample), and targeting RNAs of 17-47 nucleotides enables comprehensive assessment of the major RNA classes.

There are some limitations to JanusRNA. First, we only have serial samples from approximately 30% of the participants, so the statistical power for investigating temporal changes in cancer specific analyses is low. Also, for the control subjects there is only one sample time point available with no opportunity to measure the changes in natural variation over time. Another limitation of the dataset is that detailed clinical information from clinical registries is available only for a subset of the cases with cancer of the breast (hormone receptor status), prostate (PSA and Gleason score), lung and rectum. Pathology reports can be reviewed to complete this information (24), however that is a time-consuming process. Further, we are also missing survey data from approximately 13% of the study participants. Another weakness is the technical noise arising from analysing archived serum samples with low amounts of RNA. However, we have characterised the technical and biological variability and since technical variability is random, it has little impact on association studies.

### Reason to be cautious (limitations including generalisability)

All RNA studies, including this one, may have problems with annotation and the lack of functional information that makes the interpretation of findings challenging. Trusted annotations are essential to correctly identify transcripts, yet well-known annotation databases are not optimal (14, 25). For example, piRNA annotations contain fragments corresponding to other RNAs (26) something that might reduce comparability between trait associations. For reuse of our dataset one has to be aware of some observed batch effects correlating with changes in kit lot numbers.

## Data resource access and collaborations

The JanusRNA datasets generated for this article are not readily available because of the principles and conditions set out in articles 6 (1) (e) and 9 (2) (j) of the General Data Protection Regulation (GDPR). National legal basis as per the Regulations on population-based health surveys and ethical approval from the Norwegian Regional Committee for Medical and Health Research Ethics (REC) is also required.

There is work in progress to develop data sharing mechanisms in compliance with GDPR (27-29). In the meantime, we welcome ideas and proposals for potential collaborations for using the dataset. To facilitate this process, interested researchers can contact

## Data Availability

The JanusRNA datasets generated for this article are not readily available because of the principles and conditions set out in articles 6 (1) (e) and 9 (2) (j) of the General Data Protection Regulation (GDPR). National legal basis as per the Regulations on population-based health surveys and ethical approval from the Norwegian Regional Committee for Medical and Health Research Ethics (REC) is also required. Requests to access the datasets should be directed to the corresponding authors.

## Acknowledgements

The sequencing service was provided by the Norwegian Sequencing Centre www.sequencing.uio.no, a national technology platform hosted by Oslo University Hospital and the University of Oslo supported by the Research Council of Norway and the South-eastern Regional Health Authority. We acknowledge Dr Tom Grotmol at the CRN Department of Research and Prof Justo Lorenzo Bermejo Statistical Genetics Group, Institute of Medical Biometry & Informatics, University of Heidelberg in Germany for close collaboration and production of the testicular and gallbladder datasets, respectively. We also thank senior lawyer Hilde Olav at the CRN for help with all legal issues related to data sharing. We are thankful to the Norwegian Cancer Society for providing funding for the Janus Serum Bank management through the years of cohort recruitment. The study used data from the Cancer Registry of Norway and the Norwegian Institute of Public Health. The interpretation and reporting of these data are the sole responsibility of the authors, and no endorsement by the Cancer Registry of Norway or the Norwegian Institute of Public Health is intended nor should be inferred.

