## Supplementary Table 1 for "Data Resource Profile: thousands of circulating RNA profiles of pre-clinical samples from the Janus Serum Bank Cohort"

### Supplimentary Table of the Sequencing Data

| | Men (n=) | | | | | | | |
|---|---|---|---|---|---|---|---|---|
| Cancer site ICD 10 | Lung (C33-34) | Colon (C18) | Rectum (C19-20) | Gall bladder (C23) | Prostate (C61) | Testis (C62) | Contr - | TOT |
| No of individuals | 270 | 162 | 118 | 7 | 332 | 84 | 402 | 1375 |
| No of samples | 382 | 231 | 183 | 7 | 450 | 84 | 405 | 1742 |
| Missing data | 4 | 6 | 5 | 0 | 9 | 4 | 4 | 32 |
| RNA-seq read counts in Million mean (sd) max-min | 18.6 (6.2) 85.5-0.4 | 18.3 (5.7) 48.1-0.4 | 18.2 (5.1) 31.4-0 | 20.7 (2.9) 25.2-17.7 | 17.2 (3.6) 29.1-0.2 | 18.5 (5.3) 40.2-0.1 | 17.5 (5) 42.1-0 | 17.9 (5.2) 85.5-0 |
| miRNA counts | 284.8 (87.7) 609-103 | 285.2 (77.3) 562-119 | 294.1 (92.9) 546-2 | 292.4 (98.3) 418-169 | 287.2 (88.8) 502-21 | 347.2 (91.5) 582-2 | 294.4 (93) 568-5 | |
| isomiR counts | 2379.9 (1397.5) 9159-360 | 2278.7 (1151) 7958-518 | 2522.7 (1418.8) 8906-2 | 2065 (1205.3) 3590-638 | 2344.2 (1251.9) 7028-86 | 2952.6 (1320) 7616-2 | 2455.3 (1374.8) 8639-9 | |
| piRNA counts | 558.4 (147.5) 1216-284 | 542.9 (132) 1017-302 | 531 (141.6) 1053-25 | 511.3 (100) 676-368 | 448 (75.2) 713-108 | 568.7 (133.3) 967-21 | 531.9 (127) 1015-38 | |
| snoRNA counts | 197.6 (67.7) 484-46 | 186.1 (59.8) 408-65 | 189.3 (65.6) 378-5 | 182.1 (56.3) 268-113 | 169.6 (56.9) 396-32 | 192.2 (57.4) 315-6 | 184.5 (63.2) 436-1 | |
| lncRNA counts | 238.1 (158.2) 919-38 | 230.9 (141.6) 752-43 | 208.7 (171.8) 1041-1 | 136.1 (34.7) 166-72 | 115.6 (54.4) 355-8 | 322.6 (168.5) 733-77 | 216.1 (138.4) 838-2 | |
| mRNA counts | 2921.3 (1680.1) 8756-241 | 2786.1 (1520.8) 7986-356 | 2610.8 (1748.7) 9886-1 | 1843.1 (569.8) 2441-806 | 1484.5 (743.2) 4259-14 | 3919.9 (1411.3) 7345-2 | 2731.2 (1479.6) 8161-3 | |
| tRNA counts | 410.2 (35.7) 493-244 | 409.8 (33.7) 494-246 | 406.8 (53.5) 478-19 | 438.1 (19.7) 453-396 | 407.5 (32.4) 468-95 | 395.1 (50.9) 458-18 | 404.1 (49.6) 486-33 | |
| tRF counts | 1049.5 (439.4) 2581-175 | 1014.4 (370.4) 2450-210 | 1101.9 (461.4) 2854-3 | 937.9 (255.1) 1274-521 | 771.1 (243.2) 1565-37 | 1295 (426.5) 2148-4 | 1049.4 (446.9) 2465-8 | |

| | Women (n=) | | | | | | | |
|---|---|---|---|---|---|---|---|---|
| Cancer site ICD 10 | Lung (C33-34) | Colon (C18) | Rectum (C19-20) | Gall bladder (C23) | Breast (C50) | Ovary (C56) | Cont - | TOT |
| No of individuals | 130 | 146 | 64 | 20 | 206 | 88 | 271 | 925 |
| No of samples | 177 | 193 | 93 | 20 | 395 | 106 | 271 | 1255 |
| Missing data | 5 | 10 | 6 | 0 | 8 | 13 | 2 | 44 |
| RNA-seq read counts in Million mean (sd) max-min | 17.5 (5) 31.6-0 | 17.3 (4.9) 41.5-0.8 | 18.2 (4.8) 32.1-4.2 | 17.9 (3.7) 25.3-10.5 | 18 (3.6) 31.5-0 | 13.5 (5.9) 29.5-2.7 | 17.6 (4.3) 30.3-0.3 | 17.4 (4.6) 41.5-0 |
| miRNA counts | 272.4 (84.1) 532-25 | 288.7 (86.5) 558-36 | 275.4 (86.5) 555-105 | 257 (82.5) 432-111 | 271.3 (79.7) 515-12 | 237.3 (76.9) 415-86 | 280.1 (86.3) 502-67 | |
| isomiR counts | 2192.7 (1332.3) 9462-35 | 2431 (1433.1) 8718-92 | 2254.1 (1414.4) 8411-508 | 1917.1 (1155.9) 4465-332 | 2056.3 (1055.3) 5947-11 | 1553.8 (818.1) 4090-206 | 2210.9 (1247.2) 6483-200 | |
| piRNA counts | 539.8 (134.2) 1078-138 | 555.8 (133.2) 1246-192 | 540 (119.9) 895-340 | 433.5 (58.4) 540-337 | 419 (68) 775-34 | 383.8 (62.6) 624-262 | 481.7 (114.8) 934-97 | |
| snoRNA counts | 190.2 (66.2) 411-7 | 196.9 (58.9) 470-51 | 183.5 (62.8) 391-86 | 180.6 (68.5) 393-89 | 152.1 (46.6) 371-6 | 128.4 (42.1) 287-48 | 171.2 (58.5) 376-22 | |
| lncRNA counts | 234.3 (136.3) 829-9 | 248.8 (154.7) 889-31 | 221.3 (117.4) 615-54 | 110.2 (35.9) 199-56 | 101.6 (48.3) 330-3 | 114.3 (54.7) 317-35 | 171.6 (113.5) 698-16 | |
| mRNA counts | 2870.8 (1482.8) 7765-106 | 3046.8 (1643.7) 9859-115 | 2789.9 (1328.7) 6991-430 | 1404.8 (610.1) 2797-517 | 1289.3 (711.4) 4905-3 | 1466.1 (724) 3421-290 | 2134.7 (1383.6) 7706-51 | |
| tRNA counts | 406.7 (44.6) 490-77 | 412.7 (37.4) 490-192 | 412 (36.7) 488-297 | 425.6 (35.6) 465-351 | 406.3 (35.6) 471-36 | 364.2 (42.2) 430-216 | 407.6 (37.9) 493-71 | |
| tRF counts | 987.7 (426.4) 2537-29 | 1000.3 (400.9) 2273-35 | 1014 (439) 2319-277 | 804.3 (274.6) 1372-317 | 761.4 (227) 1598-7 | 440.5 (187.6) 934-69 | 901.7 (349.8) 2310-22 | |
